# 2D anomaly detection of fatal cerebral hemorrhage on postmortem CT

**DOI:** 10.64898/2026.09.14.26363002

**Authors:** Andrea Zirn, Eva Scheurer, Claudia Lenz

**Affiliations:** Institute of Forensic Medicine, Department of Biomedical Engineering, University of Basel, Basel, Switzerland; Institute of Forensic Medicine, Health Department Basel-Stadt, Basel, Switzerland

**Author notes:** Corresponding Author: Claudia Lenz, Institute of Forensic Medicine, Pestalozzistrasse 22, 4056 Basel, Switzerland.

**Keywords:** Anomaly detection, Diffusion models, Postmortem computed tomography, Fatal intracranial hemorrhage, Forensic imaging, Deep learning

## Abstract

Postmortem computed tomography (PMCT) is increasingly used in forensic medicine, yet image interpretation remains largely manual and time-consuming, and supervised deep learning is constrained by the scarcity of annotated forensic datasets. This study investigated whether diffusion-based generative models can detect fatal cerebral hemorrhage on PMCT through reconstruction-based anomaly detection, using solely image-level labels derived from the documented cause of death. Postmortem brain CT examinations acquired at a single forensic institute between 2011 and 2025 were retrospectively analyzed, comprising 265 non-pathological reference subjects and 33 cases of fatal intracranial hemorrhage. After automated brain extraction, slice selection and intensity normalization, two diffusion frameworks were trained: an unsupervised denoising diffusion probabilistic model (AnoDDPM) trained on non-pathological anatomy alone, and a weakly supervised classifier-guided diffusion model exploiting image-level labels. Anomaly scores were derived from the reconstruction error between each slice and its pseudo-healthy reconstruction, and detection was assessed at the image level using 95% bootstrap confidence intervals. AnoDDPM achieved an AUROC of 0.928 (95% CI 0.812-0.989) and an AUPRC of 0.794 (95% CI 0.429-0.946), whereas the classifier-guided model reached an AUROC of 0.991 (95% CI 0.984-0.997) with an AUPRC of 0.960 (95% CI 0.855-0.994). These results demonstrate excellent feasibility of diffusion-based anomaly detection for forensic PMCT and indicate that weak, image-level supervision substantially improves detection, thereby supporting the future development of automated triage tools for postmortem brain imaging.

## Introduction

Postmortem forensic imaging, often referred to as virtual autopsy or virtopsy, has significantly advanced forensic medicine by enabling non-invasive internal examinations to determine the cause of death [1]. Radiological modalities such as computed tomography (CT) and magnetic resonance imaging (MRI) are now widely used in forensic institutes either as complements or alternatives to traditional autopsies [2]. Among these modalities, postmortem CT (PMCT) is the most established due to its rapid acquisition, high spatial resolution, and ability to scan the entire body in a single session [1]. However, forensic imaging differs fundamentally from clinical imaging in both image characteristics and diagnostic goals. Postmortem changes, such as decomposition-related gas formation, lividity, and tissue desiccation, introduce artifacts that can mimic pathology [3–6]. Moreover, forensic radiologists and pathologists still spend substantial time reviewing large imaging volumes, much of which contain no relevant findings.

PMCT excels at detecting fractures, hemorrhages, gas embolism, and locating foreign bodies such as bullets or implants [7]. A study by Graziani et al. demonstrated the strength of PMCT in identifying skull fractures, brain edema, bullet trajectories, and gas distributions [8]. PMCT further offers excellent visualization of burns, heat-related injuries, and skeletal trauma [9–12], while Schober et al. demonstrated that Hounsfield unit (HU) profiles from the lungs can reflect cause-specific patterns such as cardiac arrest, hypothermia, or hemorrhage [13].

In clinical radiology, artificial intelligence (AI) has achieved expert-level performance in detecting abnormalities such as hemorrhages, fractures, and tumors, thereby motivating forensic adaptation [14–16]. For example, Ebert et al. applied convolutional neural networks (CNNs) to detect pericardial effusions on PMCT [17], and Ibanez et al. showed that CNNs trained on clinical CT scans could be fine-tuned for postmortem fractures [18]. Fehr et al. developed a pipeline based on a U-Net CNN for the automated volumetric estimation of six internal organs using PMCT data and achieved a Dice score of 0.9 compared to manual segmentation [19]. These early successes in forensic analyses highlight the feasibility and utility of AI for enhancing postmortem image interpretation [20].

Nevertheless, a major limitation is the absolute scarcity of large, annotated forensic imaging datasets. Comprehensive annotation is both time-consuming and subjective, requiring expert consensus, which hinders the scalability of supervised learning approaches. As a result, unsupervised and weakly supervised methods, such as autoencoders (AEs), variational autoencoders (VAEs), and generative adversarial networks (GANs), have gained interest in the clinical context, as these models can learn normative anatomy without requiring exhaustive labels [21–23].

More recently, diffusion models, a powerful class of generative models, have emerged as a complementary solution, surpassing GANs and VAEs in image fidelity and training stability [24–27]. Diffusion models iteratively denoise input images to generate high-quality reconstructions, enabling robust modelling of healthy anatomy and deviation-based anomaly detection [28–33]. Wolleb et al. applied diffusion-based image translation for clinical medical imaging, successfully identifying brain tumors and pleural effusions using only weak image-level labels [24]. These models excel at highlighting abnormalities by comparing the input image to a “pseudo-healthy” reconstruction and producing detailed anomaly maps. Beyond detection, the same generative principle enables high-fidelity synthetic generation of medical images, offering a route to mitigate the data scarcity that constrains forensic AI [34,35].

While this approach has not yet been validated in forensic imaging, its reliance on minimal supervision and its ability to preserve anatomical detail make it exceptionally well suited for PMCT data. Currently, no diffusion-based framework for anomaly detection in postmortem imaging exists, and there are no publicly available, large-scale, annotated datasets of PMCT scans to support training. This absence is a major bottleneck in the development of forensic AI and contributes to the continued reliance on manual interpretation. Automated anomaly detection systems could serve as triage tools and flag suspicious regions for expert review. Although fully three-dimensional approaches are ultimately desirable, initial training can leverage two-dimensional (2D) slice-wise models for efficiency and architectural maturity [36]. Intracranial hemorrhage is a leading cause of sudden death, and a previous study could demonstrate that CNNs are able to distinguish between fatal hemorrhage cases and controls on PMCT with 94 % accuracy, representing one of the first robust implementations of deep learning in forensic postmortem brain imaging [37]. Building on this foundation, the present work investigates 2D anomaly detection of cerebral hemorrhage on PMCT. As a first exploration of what can be achieved with this approach, an unsupervised and a weakly supervised diffusion framework are trained on curated reference brain datasets to establish a normative baseline of postmortem anatomy, against which deviations in fatal cerebral hemorrhage cases are detected and visualized as anomaly maps, and their performance is compared to provide a baseline for future research in this field.

## Methods

### Data collection

Postmortem brain CT examinations acquired at our Institute of Forensic Medicine between 2011 and 2025 were retrospectively reviewed. Only deceased aged 18 years or older were included. Cases were screened based on the imaging and autopsy findings, and examinations with macroscopic signs of advanced decomposition were excluded. Anomalous cases were identified based on the cause of death involving intracranial hemorrhage and comprised the full spectrum of bleedings located within the cranial cavity. Examinations without any pathological signs of the brain constituted the normal reference cohort. The raw data were stored in DICOM format as a head series with an in-plane matrix of 512 × 512 pixels and a variable number of slices along the axial (z) axis, reconstructed at a slice thickness of 0.5 mm. All examinations were acquired on one of two CT systems (Siemens SOMATOM Emotion or Siemens SOMATOM go.Now, Siemens Healthineers, Erlangen, Germany). According to national ethical standards, ethical approval was not necessary as only anonymized data were analyzed.

Image-level labels were assigned based on the documented cause of death from the autopsy records. Cases designated as normal comprised brains that showed no macroscopic signs of decomposition, no cause of death related to the brain, and no brain injuries according to the autopsy records, whereas the anomalous cohort consisted of cases in which intracranial hemorrhage constituted the cause of death and presented with visible regions of bleeding. No voxel- or pixel-wise ground-truth segmentations were employed at any stage. The final study cohort comprised 265 postmortem brain CT examinations without cerebral pathology and 33 cases with fatal intracranial hemorrhage. Across the 33 cases, a total of 71 intracranial hemorrhage findings were identified. These comprised 25 intracerebral (35%), 18 subarachnoid (25%), 15 intraventricular (21%), 12 subdural (17%), and one epidural hemorrhage (1%). Many cases showed more than one hemorrhage type.

### Data pre-processing

Since reconstruction-based anomaly detection is intended to model only the anatomy in which the pathology of interest can occur, each dataset was first reduced to a brain-only region of interest, excluding irrelevant structures and objects outside the body. The brain was automatically segmented from the surrounding skull using TotalSegmentator [38] and manually checked for accuracy. The resulting binary brain mask was multiplied voxel-wise with the original CT volume, yielding an image in which only the brain was retained. Each volume was then cropped to the brain bounding box, zero-padded to a common cubic field of view [444, 444, 444] and resampled to an isotropic resolution of 0.5 × 0.5 × 0.5 mm. Within each volume, valid axial slices were identified and slices that were blank or degraded by image artefacts were discarded. Because intracranial hemorrhage occurs at a range of anatomical levels within the brain, reference slices were sampled from the valid range to span the same distribution of anatomical levels as the hemorrhage slices. This prevented the models from discriminating slices based on anatomical position rather than the presence of pathology. For the reference volumes, 100 such axial slices were sampled per PMCT volume, whereas for the hemorrhage cases, only the slices on which the hemorrhage was visible were retained, identified by intensity thresholding within the 40 - 100 HU range characteristic of blood. Finally, a standard brain window (WC = 40 HU, WW = 80 HU) was applied to restrict the Hounsfield units to the diagnostically relevant soft-tissue range, enhancing tissue contrast, and the windowed slices were intensity-normalized to a fixed range to ensure comparability prior to model input.

### Models and training

Two complementary, state-of-the-art diffusion-based frameworks were adapted for reconstruction-based anomaly detection on PMCT. For both models, the published network architectures and training hyper-parameters were mostly retained, while the data-loading and training code was re-implemented to ensure training on our own PMCT data. All slices were processed at a resolution of 256 × 256 pixels, chosen as a compromise between anatomical detail and computational cost. All data were partitioned at the subject (volume) level, so that every slice belonging to a given individual was assigned exclusively to the training or to the held-out evaluation set and no subject appeared in more than one partition, thereby precluding information leakage between training and testing. Volumes were split at the subject level using a fixed random seed, with approximately 20 % of the volumes in each class held out as a common test set on which both models were evaluated. The remaining volumes formed the training set (reference volumes only for AnoDDPM, and both reference and hemorrhage volumes for the classifier-guided model). All models were trained and evaluated on a workstation equipped with an AMD Ryzen Threadripper 7960X CPU (24 cores) and an NVIDIA GeForce RTX 4090 GPU (24 GB).

The first model, AnoDDPM [33] is an unsupervised denoising diffusion probabilistic model trained exclusively on reference slices in order to learn a normative representation of postmortem brain anatomy. It employs a U-Net denoiser with 128 base channels, self-attention at the 32, 16 and 8 feature resolutions and two attention heads, together with a 1000-step diffusion process governed by a linear noise schedule. Following AnoDDPM’s implementation [39], corruption was adopted with multi-scale simplex noise. The network was optimized with AdamW (learning rate 1 × 10) at a batch size of one, trained for 3000 epochs with 100 iterations per epoch, and light affine augmentation (rotation ± 3°, translation up to 2 % horizonal and up to 9 % vertical shift). At inference, anomalous test slices were partially corrupted to a noise level of λ = 250 and denoised back to t = 0, similar to the original approach. The per-pixel squared difference between the input, and its reconstruction defined the anomaly map, and its mean provided the slice-level anomaly score.

The second model, the classifier-guided diffusion model of Wolleb et al. [40], is a weakly supervised approach that exploits the available image-level labels. This classifier-guided approach was implemented within the MONAI generative framework [41], using its diffusion U-Net and DDIM sampler together with a separately trained, time-step-conditioned classifier that supplies the guidance gradient. The denoising diffusion model was trained on both reference and hemorrhage slices, and a binary classifier was trained jointly to distinguish the two classes from noised images across diffusion timesteps. At inference, a test slice was encoded into noise through a sequence of deterministic DDIM steps and then decoded while its denoising trajectory was steered towards the pseudo-healthy class by the gradient of the classifier. As this guidance removes the abnormal appearance while preserving non-pathological anatomy, the difference between the input and its pseudo-healthy reconstruction localizes the pathology. The diffusion model and the classifier were trained separately using the Adam optimizer (learning rate 1 × 10) and a batch size of 10, for 1000 and 200 epochs, respectively.

The classifier-guidance scale s and the noise level L (number of DDIM encoding–decoding steps) were tuned on the validation set, as both trade off lesion removal against preservation of non-pathological anatomy. A too high L erases and mis-reconstructs normal structures, whereas a too low L leaves the hemorrhage intact. Similarly, a too small s fails to remove the lesion, while a too large s introduces artefacts. Based on this trade-off and the validation performance, L = 250 and s = 7 were selected for all subsequent experiments, as illustrated in Figure 1.

**Figure 1.**
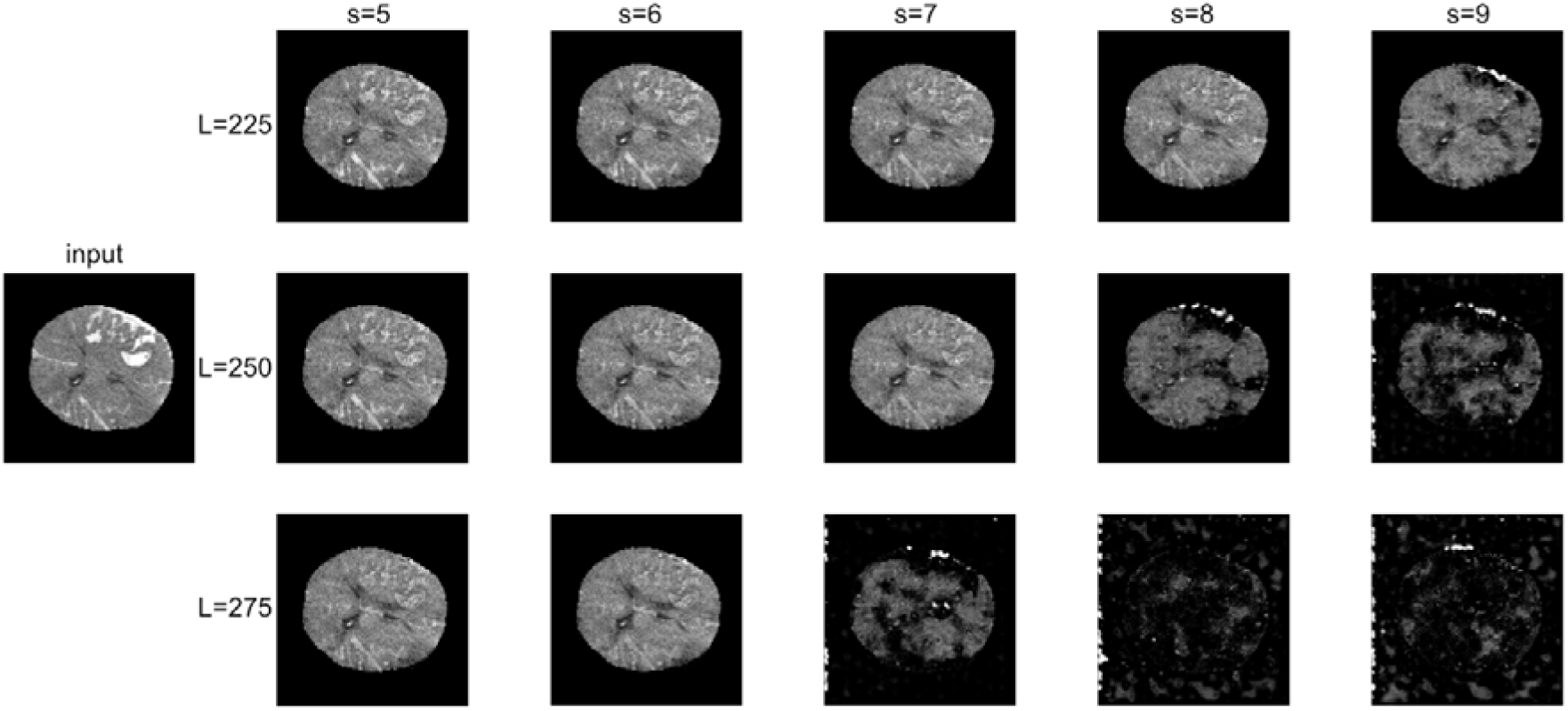
Effect of the classifier-guided inference hyperparameters on the pseudo-healthy reconstruction of a representative hemorrhage slice. Reconstructions are shown for a grid of noise levels L (rows) and classifier-guidance scales s (columns), with the input slice on the left.

### Evaluation

Because no pixel-level ground truth was available, detection performance was assessed at the image level. For each slice, the model produced a pseudo-healthy reconstruction, and the mean reconstruction error between the input and this reconstruction served as anomaly score. Reference slices, which the models reconstruct faithfully, would yield low anomaly scores, whereas slices containing hemorrhages would result in higher scores.

To account for the limited number of anomalous cases, 95 % confidence intervals for each metric were estimated by bootstrap resampling of the per-slice scores. In this procedure, the set of label-score pairs is repeatedly resampled with replacement (N = 1,000) to generate synthetic datasets of equal size, each metric is recomputed, and the 2.5^th^ and 97.5^th^ percentiles of the resulting distribution define the interval. This non-parametric approach makes no assumptions about the underlying score distribution and provides a robust estimate of variability when the sample size is small.

The choice of evaluation metric heavily depends on the anomaly detection paradigm and the granularity of detection, in our case image-level. Performance was therefore summarized by the area under the receiver-operating-characteristic curve (AUROC) and the area under the precision–recall curve (AUPRC, estimated via average precision), complemented by average precision, sensitivity, specificity, accuracy and the F1 score, the harmonic mean of precision and recall. Whereas AUROC and AUPRC are threshold-independent, the remaining measures were computed at the Youden-optimal operating point (defined as sensitivity + specificity − 1). AUROC assesses the tradeoff between true positive rate and false positive rate, offering a broad view of model discrimination. However, in highly imbalanced settings, as experienced in anomaly detection, AUROC can be overly optimistic due to its insensitivity to class distribution. In contrast, AUPRC focuses on precision and recall, making it more informative and reliable in these imbalanced scenarios by better reflecting the model’s ability to identify rare anomalies [42,43]. High sensitivity ensures anomalies are not missed while high specificity limits false alarms, which is essential when the method serves as a triage tool that flags suspicious slices for expert review. Accuracy is further reported for completeness but interpreted with caution, since under strong class imbalance, it is dominated by the majority (non-pathological) class.

In addition to these discrimination metrics, the fidelity of the pseudo-healthy reconstructions was examined by comparing the HU histograms of the reconstructed and original slices, and by comparing the apparent size of the reconstructed brain with that of the input, in order to confirm that the models reproduced normative anatomy without introducing systematic intensity or volumetric bias.

## Results

Both diffusion models were evaluated on held-out postmortem brain CT slices, each assigned an anomaly score derived from the reconstruction error. Both models were assessed on 53 reference cases (5300 slices) and 7 hemorrhage cases (489 slices). Both models separated hemorrhage from non-pathological anatomy distinctly above average, with the classifier-guided model achieving the stronger overall discrimination, as presented in Table 1 and Figure 2.

**Figure 2.**
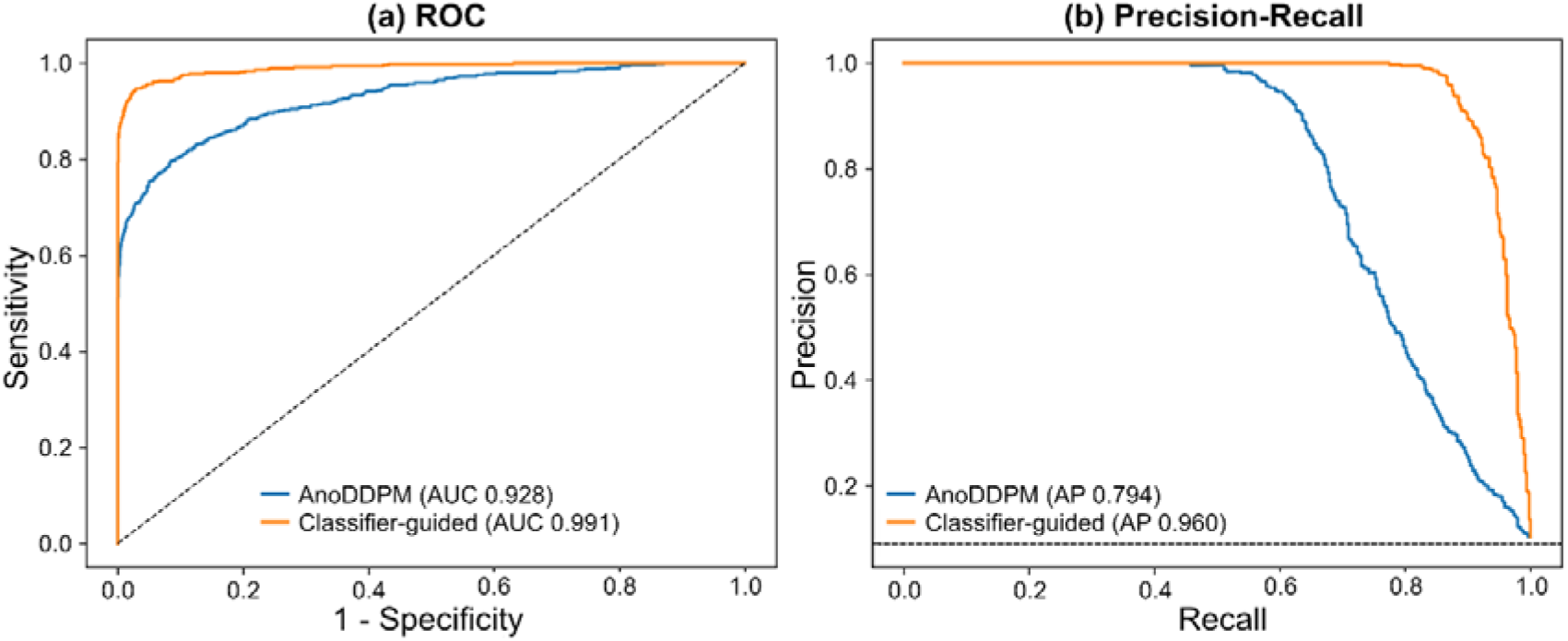
(a) Receiver-operating-characteristic and (b) precision–recall curves for the unsupervised AnoDDPM and the classifier-guided diffusion model. AUC: area under the curve; AP: average precision.

**Table 1.** Image-level (slice-wise) performance of the two diffusion models on postmortem brain CT. Confidence intervals are 95 % bootstrap intervals: sensitivity, specificity, accuracy and F1 score (*) are reported at the Youden-optimal operating point.

| **Metric** | **AnoDDPM (unsupervised)** | **Classifier-guided (weakly supervised)** |
|---|---|---|
| ROC-AUC [95 % CI] | 0.928 [0.812-0.989] | 0.991 [0.984-0.997] |
| PR-AUC [95 % CI] | 0.794 [0.429-0.946] | 0.960 [0.855-0.994] |
| Average precision | 0.794 | 0.960 |
| Sensitivity * | 0.803 | 0.991 |
| Specificity * | 0.921 | 0.962 |
| Accuracy * | 0.900 | 0.970 |
| F1 score * | 0.603 | 0.852 |

Trained on non-pathological anatomy alone, AnoDDPM, correctly identified 391 of 489 hemorrhage slices and 4876 of 5300 reference slices at the Youden-optimal operating point. The classifier-guided model, which additionally exploits image-level labels during training, achieved a higher discrimination. At the Youden-optimal threshold, it correctly detected 484 of 489 hemorrhage, missing only 5 hemorrhage slices.

The mean anomaly scores for the AnoDDPM were almost threefold higher for hemorrhage than for reference slices (0.029 vs. 0.011). For the classifier-guided approach, the mean anomaly scores were approximately 12-fold higher for hemorrhage than for reference slices (0.012 vs. 0.001). The distributions of slice-level anomaly scores, as presented in Figure 3, showed a clear upward shift for hemorrhage slices in both models, although the reference and hemorrhage ranges showed more overlapping for AnoDDPM than for the classifier-guided model.

**Figure 3.**
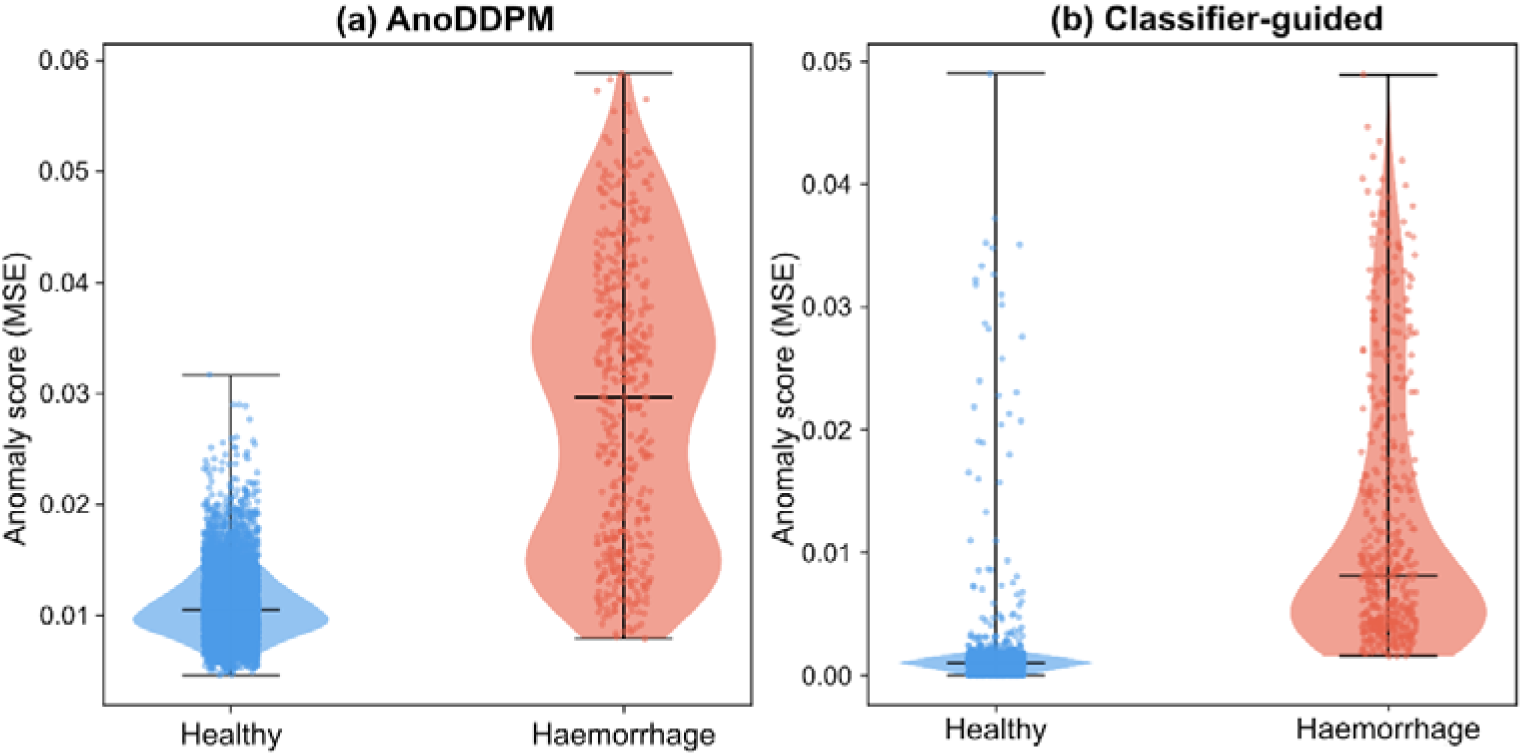
Distribution of slice-level anomaly scores for reference and hemorrhage slices for (a) AnoDDPM and (b) the classifier-guided model. Boxes show the interquartile range and median; individual slices are overlaid as points. MSE: mean square error. Note the different vertical scales.

### Qualitative assessment of anomaly maps

To complement the image-level metrics, representative pseudo-healthy reconstructions and the corresponding anomaly maps were inspected qualitatively, as shown in Figure 4. For both models, the per-pixel reconstruction error was visibly elevated over hemorrhagic regions, resulting in anomaly maps that highlighted the bleeding against a comparatively low background in non-pathological tissue. For reference cases, the anomaly maps were close to zero across the brain, reflecting the absence of pathology and supporting the specificity of the approach. The reconstructions also showed imperfect preservation of non-pathological tissue, which is further examined quantitatively in the reconstruction-fidelity analysis.

**Figure 4.**
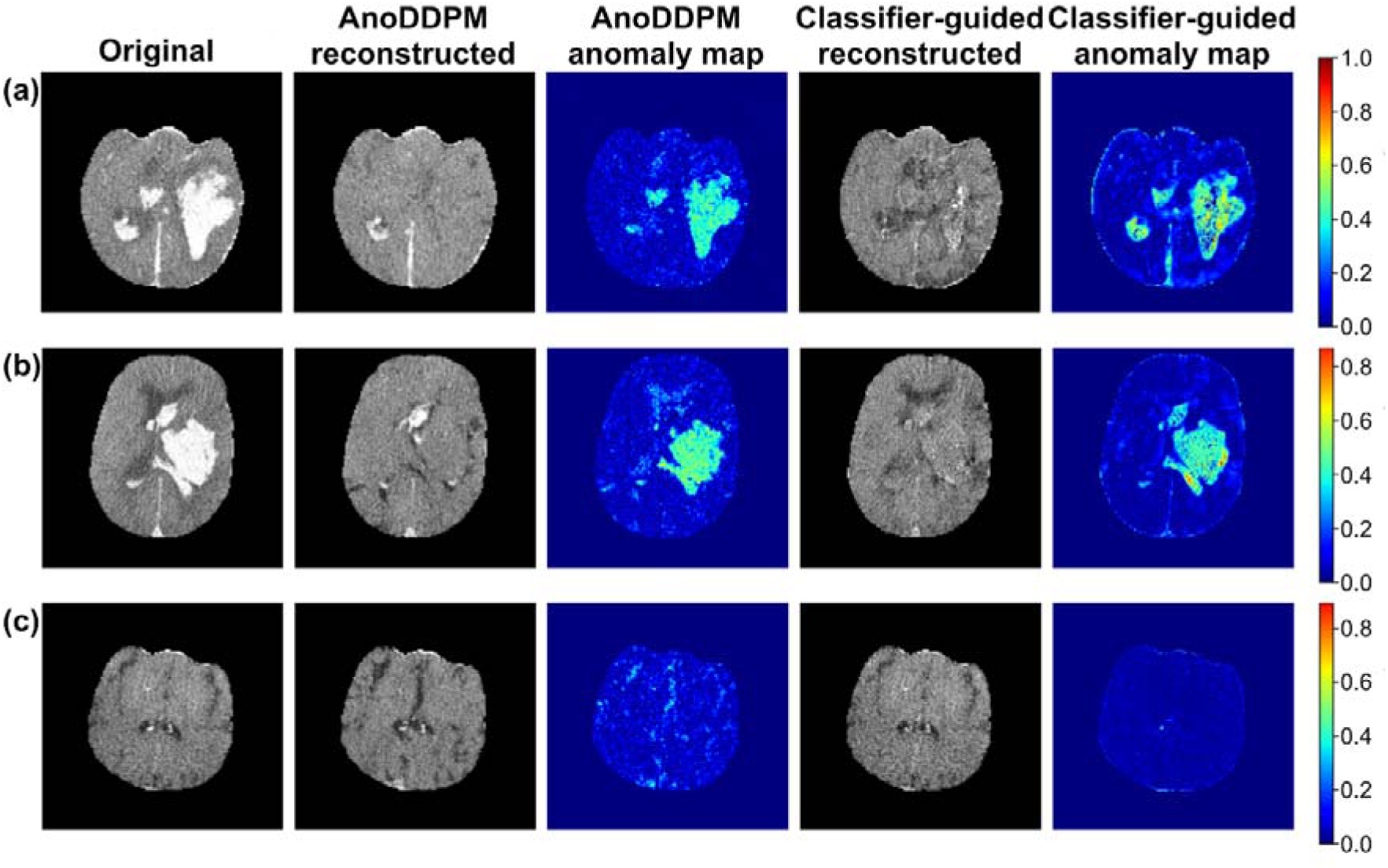
Representative results for the unsupervised AnoDDPM and the classifier-guided diffusion model. From left to right: input slice, pseudo-healthy reconstruction, and anomaly (reconstruction-error) map for AnoDDPM and classifier-guided model. Hemorrhagic regions are highlighted in the anomaly maps. (a) and (b) are example cases with fatal hemorrhage, whereas (c) represents a reference case.

### Reconstruction fidelity

Reconstruction fidelity was assessed using two complementary metrics. First, the normalized HU intensity distributions of the original and reconstructed slices were compared, as shown in Figure 5. Second, agreement between the original and reconstructed brain silhouettes was quantified using the Dice overlap and the brain-area ratio of the corresponding brain areas, as shown in Figure 6. In the HU domain, the reconstructed intensity distribution of the reference slices closely followed that of the original for both models (especially for the classifier-guided model), indicating that normative grey- and white-matter attenuation was preserved. In the HU domain (Figure 5), both models reduced the acute-blood range (≈ 55 - 75 HU) on hemorrhage slices, indicating removal of the bleeding, but the AnoDDPM also introduced a spurious peak in the normal soft-tissue range, thereby altering non-pathological tissue it should have left unchanged. For the classifier-guided model, a left shift of the overall profile can be observed, showing an incorrect reconstruction of non-pathological brain matter.

**Figure 5.**
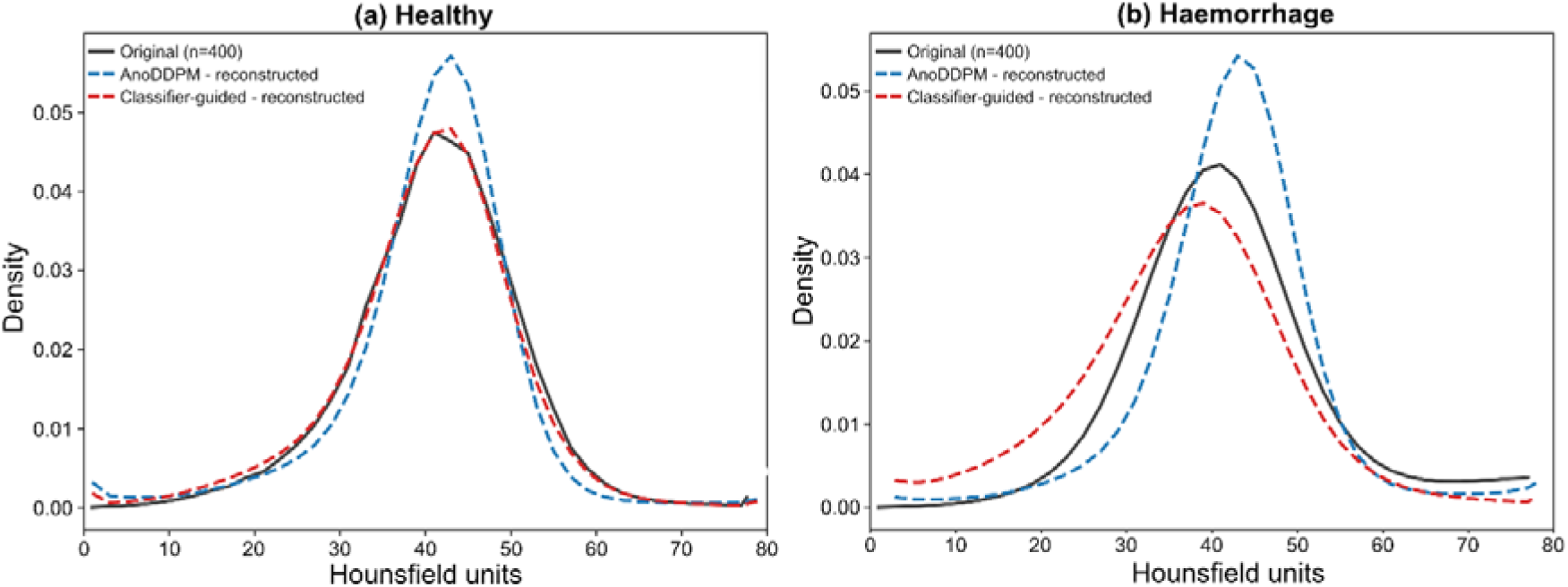
Reconstruction fidelity in the HU domain. HU intensity distributions of the original test slices (solid), the pseudo-healthy reconstructions (dashed) for AnoDDPM, as well as the classifier-guided model, shown separately for (a) reference and (b) hemorrhage slices.

**Figure 6.**
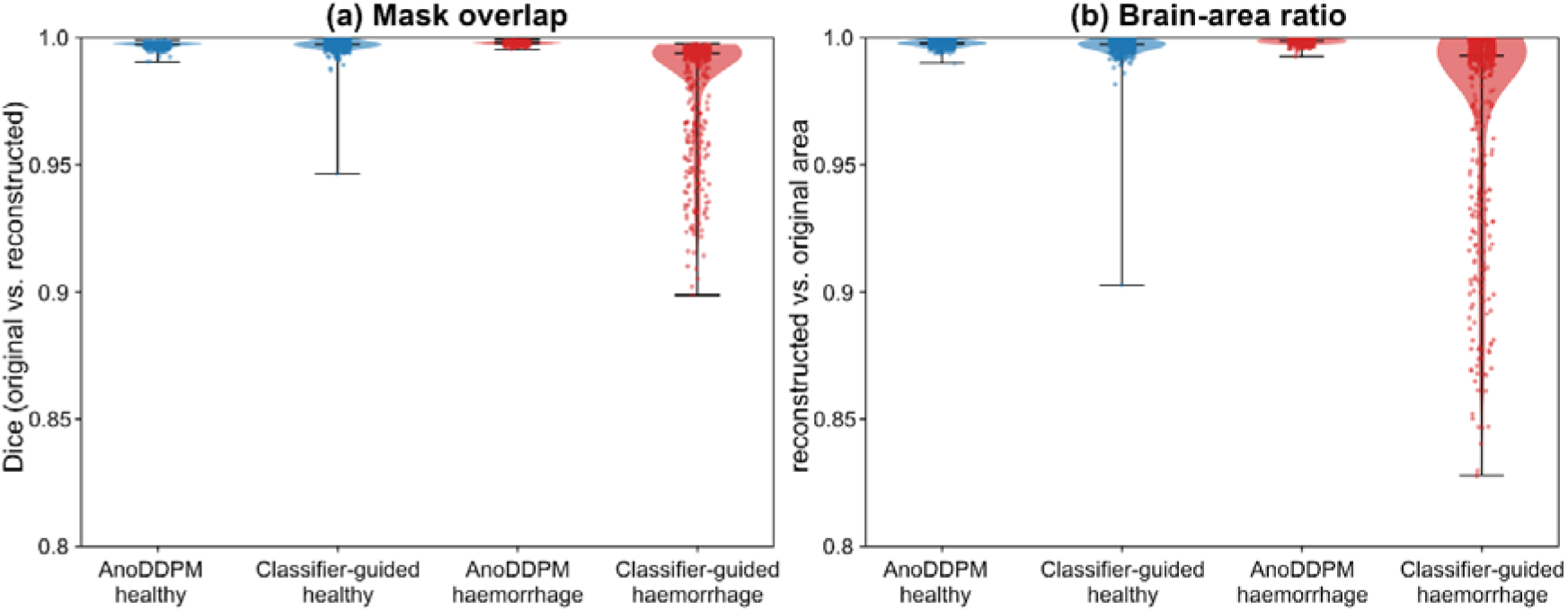
Brain-silhouette fidelity. (a) Dice overlap and (b) brain-area ratio between the original and reconstructed brain masks for AnoDDPM and the classifier-guided model across reference and hemorrhage slices. Higher values closer to 1.0 indicate more faithful preservation of the brain.

The brain silhouette analysis (Figure 6) shows that both models preserved the brain footprint almost perfectly. The original and reconstructed brain masks overlapped near-completely (Dice ≈ 0.997-0.998 for both models on reference slices and for AnoDDPM on hemorrhage slices), and the reconstructed brain area stayed within ≈ 0.2 % of the original. The only exception was the classifier-guided model on hemorrhage slices, where the mean Dice was slightly lower (0.983 ± 0.021; median 0.994) and the reconstructed area was reduced by about 2.5 %. This reflects a small number of non-ideal reconstructions under the chosen hyperparameters, which tended to erode the brain border rather than the parenchyma. Overall, both models reproduced the brain geometry faithfully, with the residual discrepancies confined to the periphery of a few hemorrhage reconstructions.

## Discussion

Our study represents the first application of diffusion-based anomaly detection to postmortem CT data. Both reconstruction-based frameworks separated hemorrhagic from non-pathological brain slices well above chance, yet their performance diverged substantially. The weakly supervised classifier-guided model of Wolleb et al. [24] reached an image-level AUROC of 0.991, markedly exceeding the 0.928 obtained by the fully unsupervised AnoDDPM [33]. This gap indicates that the coarse, image-level labels derived from the documented cause of death encode discriminative information that a purely normative model trained on non-pathological anatomy alone cannot recover, a pattern consistent with the observation that weak label guidance narrows the distance between reconstruction error and the true pathological signal [29,30].

The discrimination achieved in this work is comparable to that reported for diffusion-based anomaly detection in clinical neuroimaging, where image- and pixel-level AUROCs in the approximate range of 0.80 to 0.95 have been described for brain MRI on benchmark datasets [26,27,31,32,42,43]. Direct numerical comparison is nonetheless constrained by heterogeneity in modality, anatomy, anomaly type and evaluation granularity across these studies, a limitation emphasized by recent systematic benchmarks and reviews of medical anomaly detection [25,36,44]. Almost all prior reconstruction-based work has targeted MRI. The closest analogue to the present study is the semantic-consistent diffusion model of Sun et al. [45], which detected and segmented traumatic brain injury on cranial clinical CT and likewise demonstrated that diffusion models can localize hemorrhagic and contusional lesions without voxel-level supervision.

Several methodological choices are supported by literature. Using multi-scale simplex noise instead of independent Gaussian noise in AnoDDPM follows the original formulation [33] and accords with systematic analyses showing that the spatial scale and type of the corrupting noise govern which structures a denoising model can erase and therefore detect [46]. The classifier-guided model instead reframes detection as a counterfactual problem, generating a pseudo-healthy version of the input and attributing the residual to pathology, an approach shared by counterfactual and out-of-distribution diffusion frameworks [47,48] and subsequently refined by work on the robustness and localization accuracy of such residual maps [49,50]. The convergence of these independent publications of evidence supports the validity of the reconstruction-residual paradigm adopted here for PMCT.

Beyond the quantitative metrics, the anomaly maps provide a qualitative window onto model behavior and its limitations. In both models, the reconstruction error concentrated over hemorrhagic regions, supporting the use of the residual map as a localization cue, yet the same maps exposed two intrinsic weaknesses of the reconstruction-based paradigm. Residual error along high-contrast normative structures and a loss of fine detail introduced by the denoising process, both of which inflate the anomaly signal in non-pathological tissue and thus limit specificity. Moreover, no apparent association was observed between model performance or anomaly-map behavior and the type of intracranial hemorrhage, although the small number of cases per subtype and the frequent co-occurrence of several hemorrhage types within the same case preclude a definitive conclusion.

The reconstruction-fidelity analysis and the brain silhouette analysis further qualify this behavior. On reference slices, the reconstructions were visually faithful and free of hallucinated structures, so that the corresponding anomaly maps remained close to zero. Both models attenuated the high-attenuation blood component on hemorrhage slices, yet neither reproduced non-pathological tissue exactly. AnoDDPM introduced a spurious peak in the normal soft-tissue range while the classifier-guided model left-shifted the overall HU profile. In terms of the brain silhouette, however, both models were highly faithful, with near-complete overlap between the original and reconstructed brain masks (Dice ≈ 0.98–1.00) and reconstructed areas within a few percent of the original. The only notable exception was the classifier-guided model on hemorrhage slices (Dice 0.983 ± 0.021), where a small number of reconstructions eroded the brain border under the chosen hyperparameters. These observations are consistent with reports that the accuracy and robustness of residual-based localization depend strongly on reconstruction quality [46,49], and they motivate the future incorporation of pixel-level evaluation once reference segmentations become available.

The evaluation additionally underscores why metric selection is critical in this setting. Evaluated on the same subject-disjoint test set as AnoDDPM, the classifier-guided model attained the highest AUROC, sensitivity and F1. Because that test set is strongly imbalanced (5300 reference vs. 489 hemorrhage slices), precision and F1 remain sensitive to the reference-slice majority even at high sensitivity, which is why the area under the precision–recall curve is reported alongside AUROC. This dissociation between threshold-free ranking metrics and threshold-dependent measures under strong class imbalance is well documented and motivates the joint reporting of AUROC, AUPRC and bootstrap confidence intervals adopted here [36]. For a prospective triage application, in which a missed hemorrhage is far more costly than a false alarm later dismissed by an expert, the high sensitivity of the classifier-guided model is the more operationally relevant characteristic.

Translating these methods to forensic imaging introduces challenges absent from the clinical domain. Postmortem changes such as decomposition-related gas, hypostasis and tissue desiccation alter normative appearance and can mimic or obscure pathology, so the definition of a non-pathological postmortem brain is itself non-trivial and was here operationalized through the documented cause of death from autopsy findings rather than radiological consensus. Image-level labelling derived from the cause of death is efficient and scalable, but coarse, and may misassign individual sections on which a fatal hemorrhage is not visible. Comprehensive reviews of brain anomaly detection note that label granularity is a primary determinant of measured performance [44]. The present results nonetheless extend earlier demonstration that deep networks can distinguish fatal cerebral hemorrhage from controls on PMCT [37], advancing from binary case-level classification towards spatial localization of the bleeding.

This study has several limitations. The anomalous cohort comprised only 33 hemorrhage cases from a single institution, and although bootstrap resampling yielded narrow confidence intervals for AUROC, the wider precision-recall intervals reflect this scarcity together with the inherent class imbalance of a screening setting. Detection was quantified at the image level because no voxel-level ground truth was available. The anomaly maps were therefore assessed only qualitatively, rather than against a reference segmentation, so their spatial accuracy could not be measured. In addition, the analysis was restricted to 2D axial slices and to a single, comparatively high-contrast pathology. The 2D approach was a deliberate choice for this study: on the available hardware (a single 24 GB GPU), volumetric diffusion models would have required substantial downsampling of the PMCT volumes, sacrificing the in-plane resolution needed to depict both small and large hemorrhages. Slice-wise processing additionally yields a larger number of training samples from a limited cohort and follows the 2D formulation of the established frameworks adopted here, even at the expense of the inter-slice context that a 3D model could exploit.

Finally, only fatal intracranial hemorrhages were included in this study, whereas non-fatal or incidental bleedings were not included. Further investigation is therefore needed to determine whether the models detect smaller, non-fatal hemorrhages equally well and whether the anomaly score scales with bleeding severity, for example whether non-fatal bleedings yield lower anomaly scores than fatal ones.

These limitations outline clear directions for future work. The scarcity of annotated forensic data, the principal bottleneck identified here, may be mitigated by the same family of generative models, which have demonstrated high-fidelity synthesis of 3D medical volumes [34] and of realistic brain images through latent diffusion [35], including anatomically controllable, segmentation-guided generation that yields images paired with labels [28]. Such synthetic data, combined with cross-modality translation, for example MRI-to-CT synthesis [51], and with the use of generated labelled samples to augment segmentation training [52], could enlarge and balance training sets without additional annotation, while general-purpose synthetic-imaging models [53] and open frameworks for medical generative modelling [41] further lower the barrier to these approaches. Beyond enlarging and balancing the training data, future work should also evaluate alternative model families and architectures, for example latent-diffusion approaches, and extend the present 2D pipeline to fully volumetric 3D detection, incorporate pixel-level evaluation, and validate the system prospectively as a triage tool that flags suspect slices for expert review, which together represent the natural next steps towards forensic deployment.

## Conclusion

This work presents the first application of diffusion-based anomaly detection to postmortem CT and demonstrates that fatal cerebral hemorrhages can be localized on PMCT without voxel-level annotation. A weakly supervised classifier-guided diffusion model clearly outperformed a fully unsupervised approach, indicating that the coarse image-level labels derived from the documented cause of death provide a practical and effective form of supervision in the data-scarce forensic setting. Although validated here on a single, comparatively high-contrast pathology in two dimensions, the approach establishes a normative-deviation framework that could be extended to volumetric data, a broader range of pathologies and prospective evaluation, advancing the integration of automated image analyses into forensic postmortem practice.

## Data Availability

All data produced in the present study are available upon reasonable request to the authors.

## Acknowledgments

None

## Declaration of competing interests

None

## Notes

### Competing Interest Statement

The authors have declared no competing interest.

### Author Declarations

According to national ethical standards, ethical approval was not necessary as only fully anonymized data were analyzed.

### Summary of Updates

Improvement of text consistency regarding medical details; specifications added regarding 2D vs. 3D anomaly models.

